# Kinetics of neutralising antibodies against Omicron variant in Vietnamese healthcare workers after primary immunisation with ChAdOx1-S and booster with BNT162b2

**DOI:** 10.1101/2022.06.20.22276596

**Authors:** Nguyen Van Vinh Chau, Lam Anh Nguyet, Nguyen Thanh Dung, Vo Minh Quang, Nguyen Thanh Truong, Le Mau Toan, Le Manh Hung, Dinh Nguyen Huy Man, Dao Bach Khoa, Nguyen Thanh Phong, Nghiem My Ngoc, Huynh Phuong Thao, Dinh Thi Bich Ty, Pham Ba Thanh, Nguyen Thi Han Ny, Le Kim Thanh, Cao Thu Thuy, Nguyen To Anh, Nguyen Thi Thu Hong, Le Nguyen Truc Nhu, Lam Minh Yen, Guy Thwaites, Tran Tan Thanh, Le Van Tan, OUCRU COVID-19 Research Group

## Abstract

We studied the development and persistence of neutralising antibodies against SARS-CoV-2 ancestral strain, and Delta and Omicron (BA.1 and BA.2) variants in Vietnamese healthcare workers (HCWs) up to 15 weeks after booster vaccination. We included 47 HCWs with different pre-existing immune statuses (group 1 (G1): n=21, and group 2 (G2): n=26 without and with prior breakthrough Delta variant infection, respectively). The study participants had completed primary immunisation with ChAdOx1-S and booster vaccination with BNT162b2. Neutralising antibodies were measured using a surrogate virus neutralisation assay. Of the 21 study participants in G1, neutralising antibodies against ancestral strain, Delta variant, BA.1 and BA.2 were (almost) abolished at month 8 after the second dose, but all had detectable neutralising antibodies to the study viruses at week two post booster dose. Of the 26 study participants in G2, neutralising antibody levels to BA.1 and BA.2 were significantly higher than those to the corresponding viruses measured at week 2 post breakthrough infection and before the booster dose. At week 15 post booster vaccination, neutralising antibodies to BA.1 and BA.2 dropped significantly, with more profound changes observed in those without breakthrough Delta variant infection. Booster vaccination enhanced neutralising activities against ancestral strain and Delta variant, as compared to those induced by primary vaccination. These responses were maintained at high levels for at least 15 weeks. Our findings emphasise the importance of the first booster dose in producing cross-neutralising antibodies against Omicron variant. A second booster dose might be needed to maintain long-term protection against Omicron variant.

## INTRODUCTION

COVID-19 vaccine induced immunity wanes [1, 2], which has led to the administration of booster doses worldwide. Follow-up studies assessing the impact of booster vaccination on the development and persistence of the immune response to SARS-CoV-2 and circulating variants of concern (VOC) remain critical to informing the allocation of resources, policy decisions on COVID-19 mitigation measures, and the development of next-generation vaccines [3].

Over the last 12 months, SARS-CoV-2 Delta and Omicron VOCs have been responsible for two consecutive COVID-19 waves globally. Omicron variant is genetically divided into five major sublineages: BA.1-5. Earlier in 2022, BA.2 replaced BA.1 to become the dominant variant worldwide, including in Vietnam [4]. As of June 13^th^ 2022, BA.4 and BA.5 were responsible for the most recent waves in South Africa and Portugal [5, 6] with spread reported into Europe and the USA [7].

It is thus critical to assess levels of neutralising antibodies induced by primary course and booster vaccination against Delta and Omicron variants, especially in individuals with different pre-existing immunity; e.g. breakthrough and non-breakthrough infection. Yet, most of the reported data have been from high income countries [8-14], and few studies, especially those focusing on long term immunity, have been conducted in low- and middle-income countries.

Vietnam started the national COVID-19 vaccination programme in March 2021, and introduced the first boosters in December 2021. Herein, we focused our analysis on health care workers (HCWs) of the Hospital for Tropical Diseases (HTD) in Ho Chi Minh City (HCMC), Vietnam. Our aim was to assess the impact of the heterologous booster on the development and persistence of neutralising antibodies against the ancestral strain, Delta and Omicron variants (BA.1 and BA.2), in HTD staff with and without prior breakthrough infection.

## MATERIALS AND METHODS

### Setting and the vaccine evaluation study

The present study has been conducted at HTD in HCMC since March 2021 [15]. HTD is a 550-bed tertiary referral hospital for patients with infectious diseases in southern Vietnam. HTD has around 900 members of staff, and has been responsible for receiving COVID-19 patients of all severities in Southern Vietnam since the beginning of the pandemic.

The detailed descriptions about the study cohort have previously been reported [15]. In brief, a total of 554 individuals were enrolled at baseline, and 144 were selected for followed up from the second dose onward. Two doses of Oxford-AstraZeneca COVID-19 vaccine (ChAdOx1-S) were given as part of the primary course, completed by the first week of May 2021. And Pfizer-BioNTech COVID-19 vaccine (BNT162b2) was given as part the booster dose, completed in the third week of December 2021.

### Weekly SARS-CoV-2 testing

As per the national COVID-19 control strategy in Vietnam, between June 2021 and March 2022, HTD staff were tested weekly for SARS-CoV-2 using either PCR or antigen tests [16]. When available, samples were subjected to SARS-CoV-2 whole-genome sequencing to determine SARS-CoV-2 variant [17]. This allowed for the detection of breakthrough infection. We previously reported a cluster of breakthrough Delta variant infection among HTD staff members in June 2021 [18]. Any staff members with breakthrough infection requiring hospitalisation was admitted to HTD for clinical care.

### Plasma samples for antibody measurement

We selected 47 HCWs from the original vaccine evaluation study, consisting of group 1 (G1) including 21 without documented breakthrough infection from baseline until booster vaccination, and group 2 (G2), including 26 with breakthrough Delta variant infection [15]. More detailed descriptions about the selected participants and sampling schedules are presented in Figure 1.

**Figure 1:**
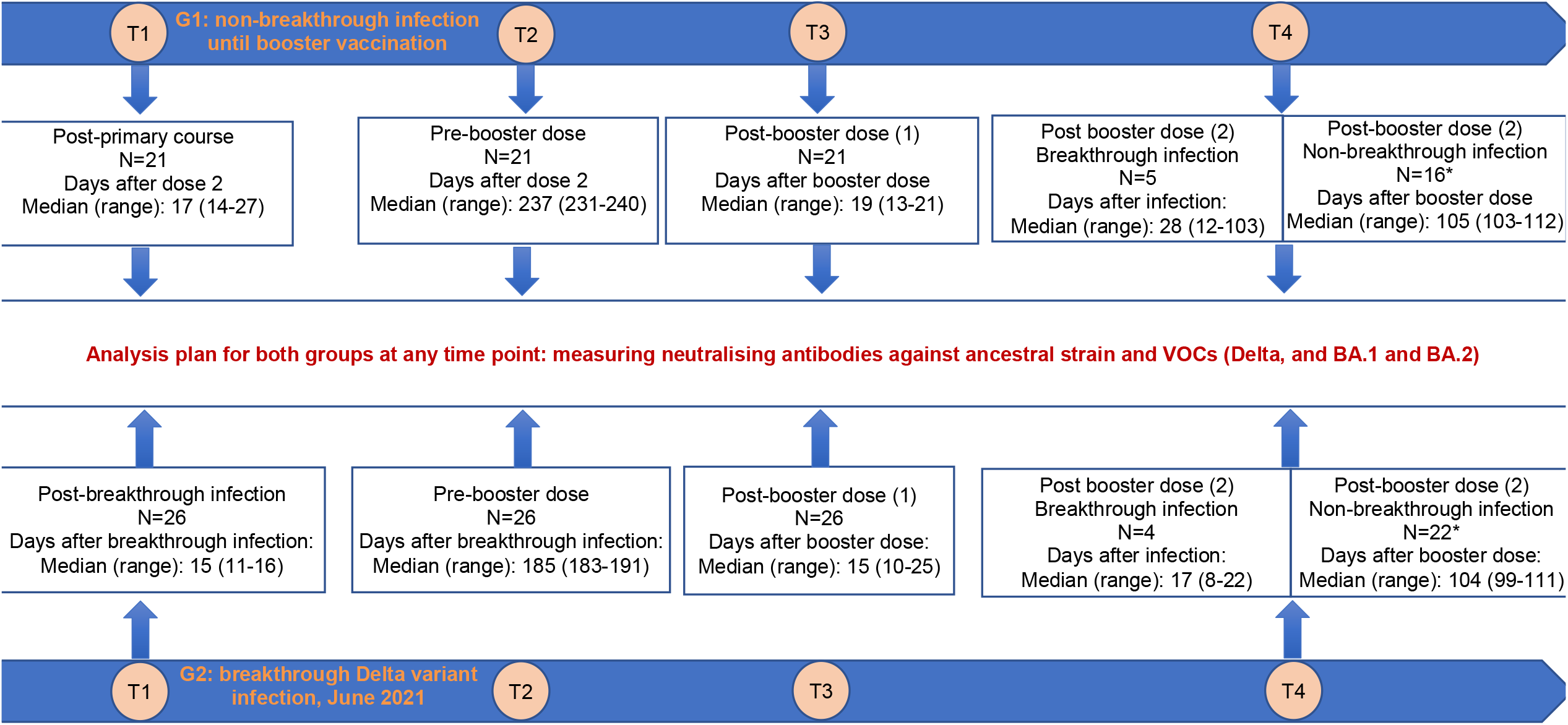
Illustration showing the distribution of the study participants and sampling schedules for neutralising antibody measurement **Note to Figure 1**: *after excluding cases with a SARS-CoV-2 infection episode recorded after the booster dose.

### Sample size justification

The 47 HCWs were selected for analysis because they had longitudinal plasma samples collected from dose 2 (G1) or breakthrough infection (G2) until month 3 after the booster dose. Sampling at this scale however has been proven to be sufficient to demonstrate the differences in antibody responses to Omicron variant in people receiving either heterologous or homologous BNT162b2 booster vaccination [13].

### Antibody measurements

For measurement of neutralizing antibodies against SARS-CoV-2 original strain (herein referred as ancestral) and SARS-CoV-2 Delta and Omicron variants (BA.1 and BA.2), we used the SARS-CoV-2 Surrogate Virus Neutralization (sVNT) assay (Genescript, USA). sVNT is a blocking ELISA that quantifies neutralizing antibodies targeting the receptor binding domain (RBD) of S protein [19]. The experiments were carried as per the manufacturers’ instructions with the readouts expressed as percentage of inhibition.

### Statistical analysis

The Wilcoxon signed-rank test or the paired T-test was used to compare the differences in neutralizing antibody levels to ancestral strain, Delta, BA.1 and BA.2 between and within groups when appropriate. The Spearman’s correlation was used to assess the correlation of neutralizing antibody levels and age. All analyses were performed using GraphPad Prism 9.3.1 (GraphPad Software, La Jolla, CA, USA).

### Ethics

The study received approvals from the Institutional Review Board of the Hospital for Tropical Diseases in Ho Chi Minh City Vietnam and the Oxford Tropical Research Ethics Committee. Written informed consent was obtained from all the study participants.

## RESULTS

### Demographics and breakthrough infection after booster vaccination

Information about the demographics and vaccination status of the selected participants are presented in Table 1 and Figure 1. The window time between the second dose and the booster dose was around 8 months. Of the 26 participants in G2, the window time from infection to booster vaccination was around 6 months (Figure 1).

**Table 1:** Demographics and time intervals between vaccine doses **Note to Table 1:** NA: Non-applicable

| Variables | G1: HCWs without documented breakthrough infection prior to booster vaccination (n=21) | G2: HCWs with breakthrough infection prior to booster vaccination, (n=26) |
| --- | --- | --- |
| Male gender, n (%) | 1 (4.8) | 10 (38.5) |
| Age year, median (range) | 35 (24-54) | 40.5 (24-56) |
| Vaccine dose 1 date (range) | 8-12 Mar/2021 | 8-15 Mar/2021 |
| Vaccine dose 2 date (range) | 19-28 Apr/2021 | 22 Apr - 4 May/2021 |
| Vaccine dose 3 date (range) | 16-17 Dec/2021 | 16-21 Dec/2021 |
| Days from vaccine dose 1 to dose 2, median (range) | 43 (40-49) | 44 (39-53) |
| Days from vaccine dose 2 to dose 3, median (range) | 238 (233-241) | 238 (231-245) |
| Days from vaccine dose 3 to breakthrough infection, median (range) | NA | 187 (184-191) |
| Breakthrough infection after booster dose, n (%) | 5 (23.8) | 4 (15.4) |
| Days from booster vaccination to infection, median (range) | 82 (1-94) | 86 (77-96) |
| Days from infection after booster dose to blood sampling, median (range) | 28 (12-103) | 17 (8-22) |

**Note to Table 1:** NA: Non-applicable

During the follow up, 9 individuals, including 5/21 (24%) participants of G1 and 4/26 (15%) participants of G2, had a SARS-CoV-2 infection episode recorded after the booster dose (Figure 1). Although detailed clinical descriptions were not available, no hospitalisation was reported, suggesting that all were either asymptomatic or mildly symptomatic. E-gene real time PCR Ct values were available in two samples of G1 (13.1 and 15.4). Of these, information about SARS-CoV-2 variant was available in one; which was assigned to BA.2. The window time (median in days) from infection to blood sampling at month three after booster vaccination was 28 (range: 12-103) for G1 and 17 (range: 8-22) for G2.

### Neutralising antibodies against BA.1 and BA.2 after primary vaccination with ChAdOx1-S

Of the 21 participants in G1, at week two after the primary course, detectable neutralising antibodies against ancestral strain and Delta variant were documented in 21 (100%) and 20 (95%), with comparable levels to ancestral strain and Delta variant (Figure 2A and Table 2). Neutralising antibodies to BA.2 were not detected, while neutralising antibodies against BA.1 were detected in only 3 participants but the titers approached the detection limit of the sVNT assay (Figure 2A and Table 2).

**Table 2:** Neutralising antibody levels to ancestral strain, Delta variant, BA.1 and BA.2 measured at four time points during the study period **Note to Table 2:** Reported values are median inhibition in % (interquartile range)

|  |  | Ancestral strain | Delta variant | BA.1 | BA.2 |
| --- | --- | --- | --- | --- | --- |
| G1: HCWs without prior breakthrough infection | 2 weeks after dose 2 | 86.2 (74.6-93.8) | 86.3 (74.4-89.8) | -4.7 (-12-9.5) | 6.1 2.9-14.4) |
|  | Before dose 3 | 26.9 (19.1-36.2) | 7.5 (-5.3-17.5) | -3.2 (-5.5-1) | 0 (0-3.3) |
|  | 2 weeks after dose 3 | 97.3 (96.9-97.5) | 98.2 (98.0-98.3) | 84.1 (74.1-90.6) | 92.7 (89.2-95.6) |
|  | 15 weeks after dose 3 | 97.5 (97.4-97.6) | 97.9 (97.4-98.1) | 54.3 (19.1-92.1) | 83.7 (68.3-97.6) |
| G2: HCWs with prior breakthrough infection | 2 weeks after breakthrough infection | 96.9 (81.6-97.0) | 97.6 (77.2-98.2) | 70.0 (10.9-80.8) | 85.7 (-5.2-92.4) |
|  | Before dose 3 | 96.0 (87.8-96.6) | 97.5 (71.9-98) | 18.8 (6-46.7) | 74.1 (34.5-82.9) |
|  | 2 weeks after dose 3 | 96.1 (95.8-97.5) | 98.1 (98.0-98.4) | 92.3 (82.8-95.7) | 95.7 (90.8-97.7) |
|  | 15 weeks after dose 3 | 96.8 (96.7-97.0) | 98 (98.0-98.2) | 87.8 (70.5-93.2) | 94.9 (89.0-98.2) |

**Figure 2:**
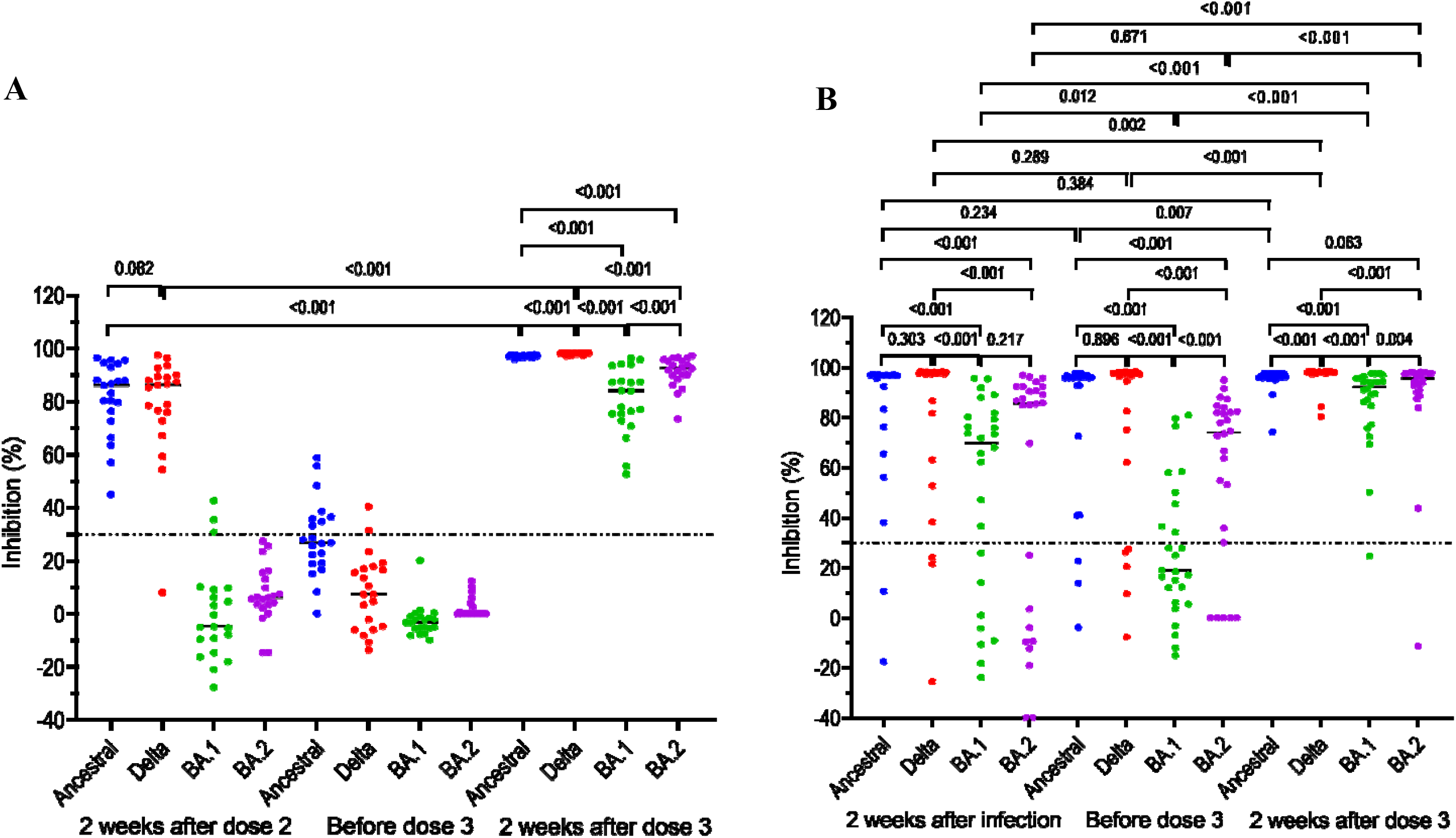
Neutralizing antibodies against SARS-CoV-2 ancestral strain and VOCs (Delta, BA.1 and BA.2) in individuals with and without prior breakthrough Delta variant infection measured at different time points prior to booster vaccination and at week 2 post booster dose. Horizontal dot lines indicate assay cut-off. Numbers indicates p values.

### Development of neutralising antibodies after heterologous booster with BNT162b2 in individual with and without prior breakthrough infection

Of the 21 participants in G1, before the booster dose (i.e. month 8 after dose 2), none had detectable neutralising antibodies against Omicron variant (BA.1 and BA.2). The proportions of individuals with detectable neutralising antibodies to the ancestral strain and Delta variant were 8/21 (38%) and 2/21 (10%), respectively, with neutralising titers approaching the assay detection limit (Figure 2A). At week 2 after the booster dose, all had neutralising antibodies against ancestral strain and VOCs (Delta, BA.1 and BA.2). Notably neutralising antibody levels to ancestral strain, and Delta variant measured at 2 weeks after the booster dose were significantly higher than those to the respective viruses measured at week 2 after dose 2 (median inhibition in % (interquartile range (IQR)): for ancestral strain: 97.3 (96.9-97.5) vs. 86.2 (74.6-93.8), p<0.001, and for Delta variant: 98.2 (98.0-98.3) vs. 86.3 (74.4-89.8), p<0.001) (Figure 2A). Neutralising antibody levels to BA.2 were significantly higher than those to BA.1 (median inhibition in % (IQR): 92.7 (89.2-95.6) vs. 84.1 (74.1-90.6), p<0.001) (Figure 2A and Table 2).

Of the 26 participants in G2, neutralising antibodies against ancestral strain, Delta, BA.1 and BA.2 measured at week 2 after breakthrough infection were detectable in 24 (92%), 23 (88%), 18 (69%) and 17 (65%), respectively (Figure 2B), with neutralising antibody levels to the ancestral strain and Delta variant significantly higher than those to BA.1 and BA.2 (Figure 2B and Table 2). At week 2 after booster vaccination, neutralising antibody levels to BA.1 and BA.2 significantly increased as compared to those measured before the booster dose and at 2 weeks post breakthrough infection, but remained significantly lower than those against ancestral strain and Delta variant (Figure 2B and Table 2). At this time point, neutralising antibody levels to BA.2 were significantly higher than those to BA.1 (median inhibition in % (IQR): 95.7 (90.8-97.7) vs. 92.3 (82.8-95.7), p<0.004) (Figure 2B).

### Persistence of neutralising antibodies at week 15 after booster vaccination

To assess the persistence of neutralising antibodies induced by the booster dose, we first focused our analysis on those without a SARS-CoV-2 infection episode documented after booster vaccination. At week 15 after the booster dose, of 16 study participants in G1, 16 (100%) had detectable neutralising antibodies against ancestral, Delta and BA.2 variants, while 11/16 (69%) had detectable neutralising antibodies against BA.1. Accordingly, neutralising antibody levels to BA.1 and BA.2 was significantly lower compared to those measured at week 2 post booster vaccination (median inhibition in % (IQR): for BA.1: 54.3 (19.1-92.1) vs. 85.7 (71.3-92.1), p=0.034, and for BA.2: 83.7 (68.3-97.6) vs. 93.2 (90.4-95.7), p=0.034) (Figure 3A). Neutralising antibodies against Delta variant also slightly reduced, but remained at very high titers (Figure 3A).

**Figure 3:**
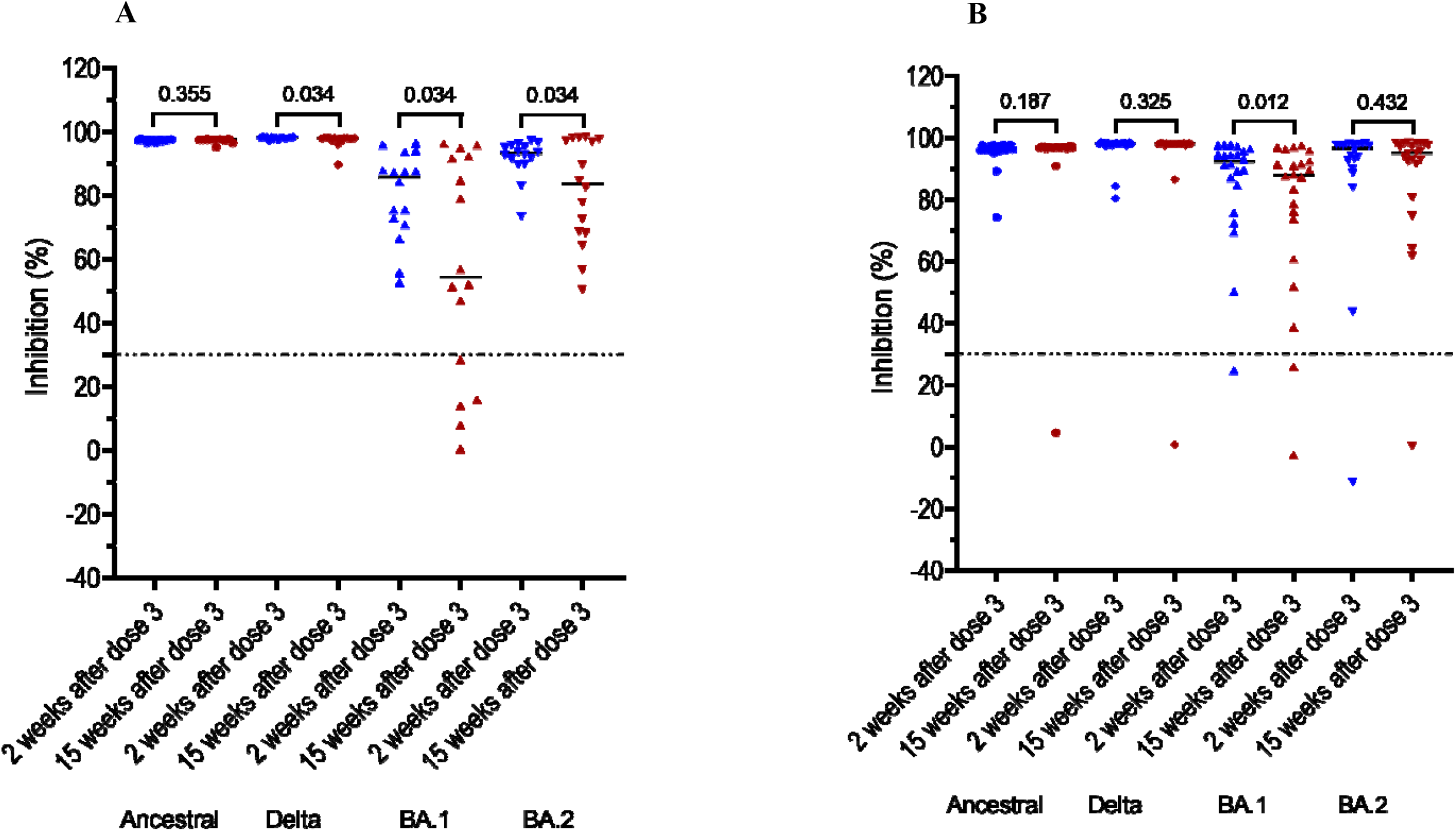
Persistence of neutralising antibodies at month 3 after the booster dose in those without documented breakthrough infection after the booster dose, **A**) participants of G1, and **B**) participants of G2. Horizontal dot lines indicate assay cut-off, Numbers indicates p values.

Of the 22 participants without a SARS-CoV-2 infection episode documented after the booster dose in G2, neutralising antibody levels to BA.1 significantly decreased (median inhibition in % (IQR): 87.8 (70.5-93.2) vs. 92.3 (82.5-95.8), p=0.012) (Figure 3B). Otherwise, neutralising antibody levels to ancestral strain, Delta and BA.2 measured at this time points were comparable with those of the corresponding viruses measured at two weeks post booster dose (Figure 3B and Table 2).

Of the nine individuals with a documented SARS-COV-2 infection episode after the booster dose, neutralising antibody levels to ancestral strain and all VOCs slightly increased at week 15, albeit not statistically significant in case of ancestral strain, BA.1 and BA.2 (Supplementary Figure 1).

### Association between age and neutralising antibody levels to BA.1 and BA.2

Results of linear regression analysis showed no association between age and neutralizing antibodies levels to BA.1 and BA.2 measured at week 2 and week 15 post booster dose (Supplementary Figure 2). Similar analysis for ancestral strains and Delta variant was considered uninformative because neutralising antibody levels to these two viruses in all study participants reached the upper detection limit of the assay (100%) (Figure 3).

## DISCUSSION

We showed that neutralising antibodies induced by primary immunization with ChAdOx1-S in Vietnamese HCWs failed to neutralize Omicron variant BA.1 and BA.2. Heterologous booster vaccination with BNT162b2 improved the immunity that could broadly neutralize both BA.1 and BA.2 in HCWs with and without prior breakthrough infection. Additionally, booster vaccination significantly enhanced neutralising antibody levels to the ancestral strain and Delta variant. However, neutralising antibodies against BA.1 and BA.2 significantly declined at month 3 post-booster vaccination, particularly in those without breakthrough infection, while neutralising antibodies to ancestral strain and Delta variant remained at high titers. We found no association between age and neutralising antibody levels, in line with a recent report [12], but none of our study participants were older than 57 years. Our findings are consistent with existing data regarding the capacity of the Omicron variant to escape from neutralizing antibodies induced by vaccination [10, 20]. The results also support previous findings about the effectiveness of the third doses in preventing infection, severe disease and death [14]. Over half of the plasma samples collected at two weeks after breakthrough Delta variant infection cross-neutralised BA.1 and BA.2, supporting recent reports regarding protection against Omicron offered by previous infection [21-23]. Booster vaccination further enhanced the cross-neutralising activities and the proportion of plasma samples with detectable neutralizing antibodies in these individuals with breakthrough Delta variant infection 6 months earlier [18]. Because neutralising antibodies titers are well correlated with protection [24, 25], the data suggest that booster vaccination could still be beneficial to individuals with breakthrough infection in protecting against Omicron variant [21, 23]. Likewise, the decline in neutralsing antibody levels to sublineages BA.1 and BA.2 at week 15 after the first booster dose suggest that a second booster doses might be needed to maintain the long-term protection of vaccine against Omicron variant [12]. Our results also compliment findings from a recent population based study in the USA [2], which showed that during the Omicron wave vaccine effectiveness against hospitalizations dropped from 91% during the first 2 months to 78% ≥4 months after a third dose. Additionally, a recent study from Israel demonstrated that a second booster dose of the BNT162b2 vaccine was effective in reducing the risk of COVID-19 associated outcomes (including infection) in individuals already completing the first booster dose at least 4 months earlier [9].

Our study consistently showed that neutralising antibody tiers against BA.2 after the booster dose in individuals with and without prior breakthrough infection were significantly higher than those against BA.1. Relevant data from previous studies have so far been inconsistent. Recent studies from Germany and Hong Kong showed comparable serum neutralising antibody levels to BA.1 and BA.2 in individuals completing three doses of BNT162b2 [8, 26]. In contrast, Yu and colleagues showed that median neutralising antibody titers against BA.2 was lower than those against BA.1 in people triple vaccinated with BNT162b2, and in those with previous infection regardless of the vaccination status [11]. The differences in study populations and pre-existing immunity induced by past exposure and/or vaccination might be contributing factors. Whether BA.2 is less able to evade immunity than BA.1 merits further research.

Our study has several limitations. First, we did not perform live virus neutralisation assay, currently the gold standard, to measure neutralising antibodies. However, the percentage of inhibition measured by the sVNT test has been shown to correlate well with the neutralizing antibody titers measured by the conventional plaque reduction neutralization assay [19]. Second, we did not study T-cell responses, which have been proven to play an important role in protecting against severe disease and death, and in case of Omicron variant, despite the neutralisation escape, T-cell responses were preserved at around 70-80% [27, 28].

In summary, we showed that booster vaccination by BNT162b2 induced cross-neutralising activities against sublineages BA.1 and BA.2 of Omicron variant in Vietnamese HCWs completing primary immunization with ChAdOx1-S. These responses however significantly reduced at month 3 post booster doses, indicating a second booster is potentially needed to maintain long-term vaccine effectiveness against the currently circulating variants. Vaccines remains critical to reduce the transmission and to protect against severe disease and death.

## Data Availability

All data produced in the present work are contained in the manuscript.

## ACKNOWLEDGEMENTS

This study was funded by the Wellcome Trust of Great Britain (106680/B/14/Z, 204904/Z/16/Z and 222574/Z/21/Z).

We thank our colleagues at the Hospital for Tropical Diseases in Ho Chi Minh City, Vietnam, for their participations in this study.

OUCRU Vietnam COVID-19 research group: Chambers Mary, Choisy Marc, Day Jeremy, Dong Huu Khanh Trinh, Dong Thi Hoai Tam, Du Hong Duc, Dung Vu Tien Viet, Fisher Jaom, Flower Barney, Geskus Ronald, Hang Vu Thi Kim, Ho Quang Chanh, Ho Thi Bich Hai, Ho Van Hien, Hung Vu Bao, Huong Dang Thao, Huynh le Anh Huy, Huynh Ngan Ha, Huynh Trung Trieu, Huynh Xuan Yen, Kestelyn Evelyne, Kesteman Thomas, Lam Anh Nguyet, Lawson Katrina, Leigh Jones, Le Kim Thanh, Le Dinh Van Khoa, Le Thanh Hoang Nhat, Le Van Tan, Lewycka Sonia Odette, Lam Minh Yen, Le Nguyen Truc Nhu, Le Thi Hoang Lan, Nam Vinh Nguyen, Ngo Thi Hoa, Nguyen Bao Tran, Nguyen Duc Manh, Nguyen Hoang Yen, Nguyen Le Thao My, Nguyen Minh Nguyet, Nguyen To Anh, Nguyen Thanh Ha, Nguyen Than Ha Quyen, Nguyen Thanh Ngoc, Nguyen Thanh Thuy Nhien, Nguyen Thi Han Ny, Nguyen Thi Hong Thuong, Nguyen Thi Hong Yen, Nguyen Thi Huyen Trang, Nguyen Thi Kim Ngoc, Nguyen Thi Kim Tuyen, Nguyen Thi Ngoc Diep, Nguyen Thi Phuong Dung, Nguyen Thi Tam, Nguyen Thi Thu Hong, Nguyen Thu Trang, Nguyen Thuy Thuong Thuong, Nguyen Xuan Truong, Nhung Doan Phuong, Ninh Thi Thanh Van, Ong Phuc Thinh, Pham Ngoc Thanh, Phan Nguyen Quoc Khanh, Phung Ho Thi Kim, Phung Khanh Lam, Phung Le Kim Yen, Phung Tran Huy Nhat, Rahman Motiur, Thuong Nguyen Thi Huyen, Thwaites Guy, Thwaites Louise, Tran Bang Huyen, Tran Dong Thai Han, Tran Kim Van Anh, Tran Minh Hien, Tran Phuong Thao, Tran Tan Thanh, Tran Thi Bich Ngoc, Tran Thi Hang, Tran Tinh Hien, Trinh Son Tung, van Doorn H. Rogier, Van Nuil Jennifer, Vidaillac Celine Pascale, Vu Thi Ngoc Bich, Vu Thi Ty Hang, Yacoub Sophie. HTD COVID-19 research group: Nguyen Van Vinh Chau, Nguyen Thanh Dung, Le Manh Hung, Huynh Thi Loan, Nguyen Thanh Truong, Nguyen Thanh Phong, Dinh Nguyen Huy Man, Nguyen Van Hao, Duong Bich Thuy, Nghiem My Ngoc, Nguyen Phu Huong Lan, Pham Thi Ngoc Thoa, Tran Nguyen Phuong Thao, Tran Thi Lan Phuong, Le Thi Tam Uyen, Tran Thi Thanh Tam, Bui Thi Ton That, Huynh Kim Nhung, Ngo Tan Tai, Tran Nguyen Hoang Tu, Vo Trong Vuong, Dinh Thi Bich Ty, Le Thi Dung, Thai Lam Uyen, Nguyen Thi My Tien, Ho Thi Thu Thao, Nguyen Ngoc Thao, Huynh Ngoc Thien Vuong, Huynh Trung Trieu, Pham Ngoc Phuong Thao, Phan Minh Phuong. EOCRU COVID-19 research group: Bachtiar, Andy, Baird, Kevin J., Dewi, Fitri, Dien, Ragil, Djaafara, Bimandra A., Elyazar, Iqbal E., Hamers, Raph L., Handayani, Winahyu, Kurniawan, Livia Nathania, Limato, Ralalicia, Natasha, Cindy, Nuraeni, Nunung, Puspatriani, Khairunisa, Rahadjani, Mutia, Rimainar, Atika, Saraswati, Shankar, Anuraj H., Suhendra, Henry, Sutrisni, Ida Ayu, Suwarti, Tarino, Nicolas, Timoria, Diana, Wulandari, Fitri. OUCRU-NP COVID-19 research group: Basnyat Buddha, Duwal Manish, Gautum Amit, Karkey Abhilasha, Kharel Niharika, Pandey Aakriti, Rijal Samita, Shrestha Suchita, Thapa Pratibha, Udas Summita.

## Supplementary Materials For

**Supplementary Figure 1:**
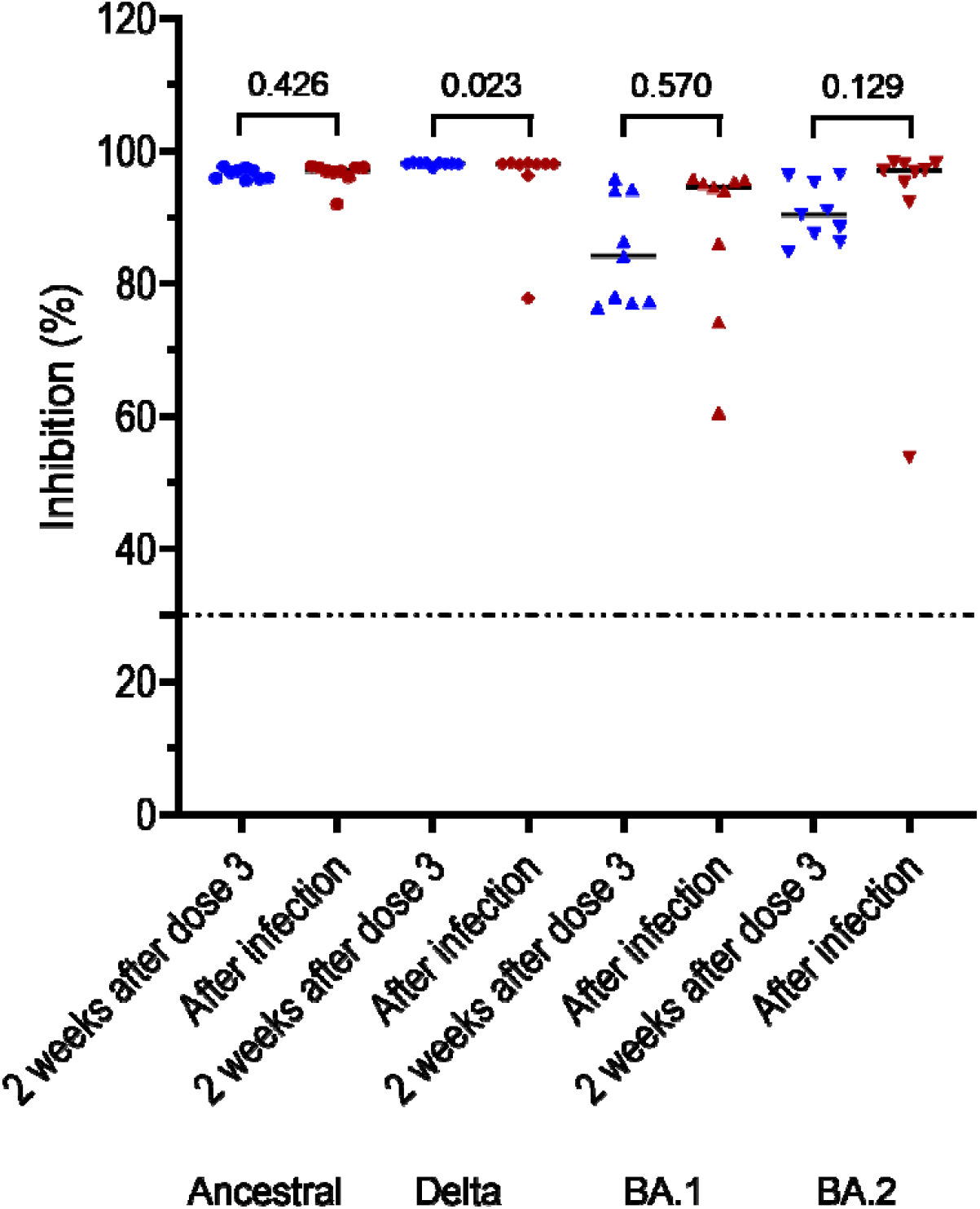
Neutralising antibodies measured at week 2 and 15 after booster vaccination in 11 HCWs with an infection episode documented after the booster dose

**Supplementary Figure 2:**
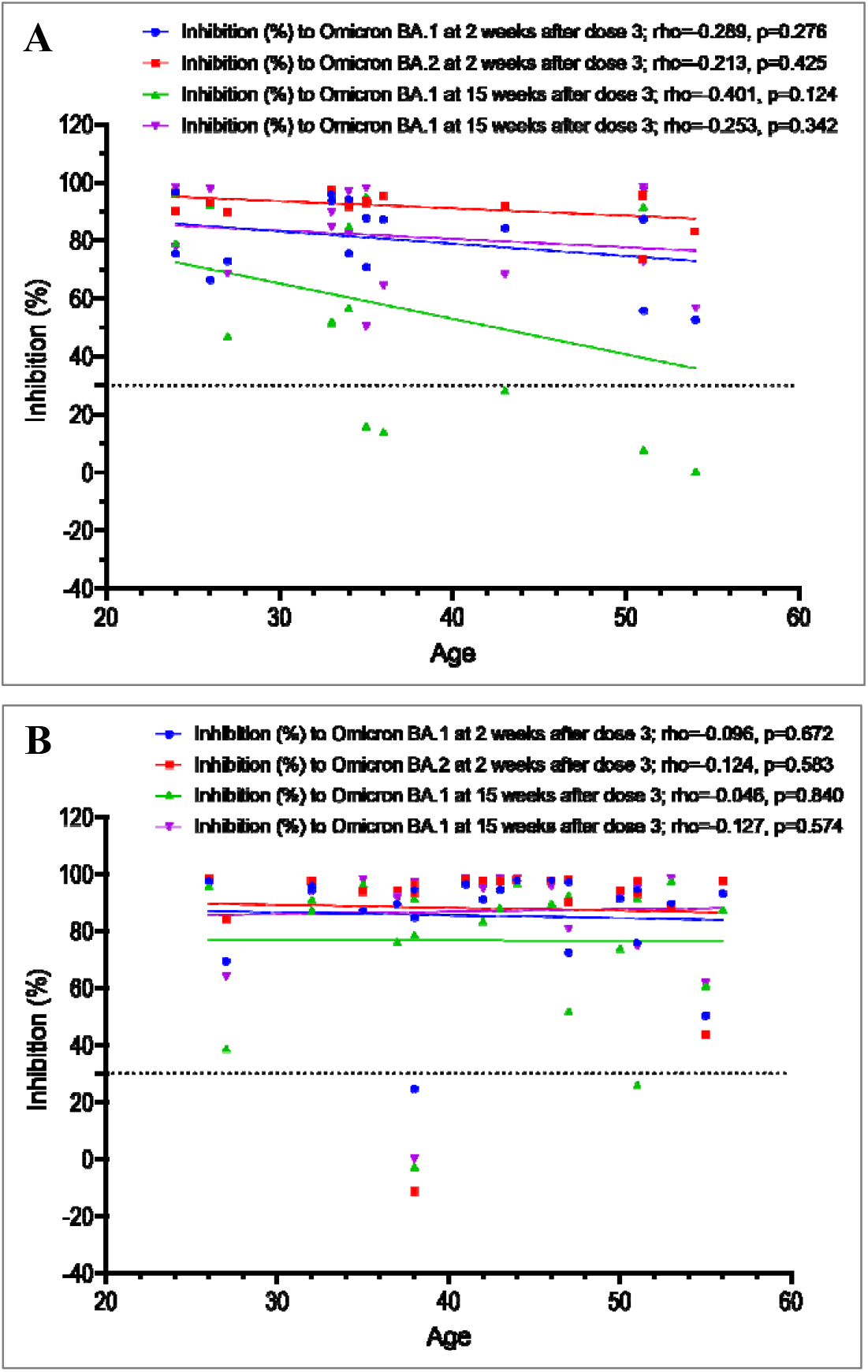
Association between age and neutralising antibody levels to BA.1 and BA.2 measured at week 2 and 15 post booster vaccination in those without a SARS-CoV-2 infection episode recorded after the booster dose, A): 16 individuals of G1 and B) 22 individuals of G2 **Note to Supplementary Figure 2**: rho: Spearman’s rank correation coefficient

